# Association between Blood Lead Levels and Anaemia in a Pregnant Population in the Kintampo Area of Ghana: A Prospective Cohort Study

**DOI:** 10.64898/2026.10.23.26366301

**Authors:** Veronica Agyemang, Otchere Addai-Mensah, Lilian Antwi Boateng, Benedict Sackey, David Dosoo, Dennis Adu-Gyasi, Dennis Gyasi Konadu, Irene Apewe Adjei, Lawrence Febir, Stephaney Gyaase, Joana Edem Koto, Eliezer Ofori-Lartey, Charlotte Tawiah Agyemang, Emily R. Smith, Sam Newton, Kwaku Poku Asante

## Abstract

**Background:** Maternal anaemia and lead exposure both pose substantial risks in pregnancy, but their relationship has not been consistently explored. We assessed trimester-specific blood lead levels (BLLs) and their association with anaemia in pregnant women in the Kintampo area of Ghana.

**Methods:** In this prospective cohort study nested within the PRISMA Maternal and Newborn Health Study in Ghana, 300 pregnant women were enrolled at <14 weeks’ gestation and followed until delivery. Trimester-specific venous blood was collected and analysed for haemoglobin levels using Sysmex XN 330 auto analyser (Sysmex Corporation, Japan) and BLL using inductively coupled plasma-mass spectrometry (ICP-MS). Anaemia was defined as 11.0g/dL for first (T1) and third (T3) trimesters and 10.5g/dL for second (T2) trimester according to the World Health Organisation’s (WHO’s) trimester-specific haemoglobin thresholds and elevated BLL as ≥3.5 µg/dL by the Centers for Disease Control and Prevention (CDC). Multiple logistic regression, adjusted for maternal age, parity, education, malaria parasitaemia, body mass index, gestational age and occupation, examined the BLL and anaemia association. A population-averaged Generalized Estimating Equations (GEE) model was used to assess the association between elevated BLL and anaemia across trimesters.

**Results:** The mean age of participants was 27.7±5.8 years (range 18-44years). Geometric mean BLL increased significantly across trimesters from 1.0 to 1.3 to 1.5 µg/dL (*p*=0.003) for T1, T2 and T3 respectively. The prevalence of elevated BLL (≥3.5 µg/dL) was 10.7%, 7.6%, and 15.2% for T1, T2, and T3 respectively. Anaemia prevalence increased significantly with increasing trimester from 34.7.0% to 59.3% to 76.0% (p<0.001) for T1, T2 and T3 respectively. In the population-averaged GEE model, elevated BLL was associated with increased odds of anaemia across trimesters, though this did not reach statistical significance (OR = 1.29, 95% CI: 0.77–2.17, p = 0.327).

**Conclusion:** Anaemia prevalence in pregnancy is high in the Kintampo area and increases with advancing gestation. No significant association was observed between BLL and anaemia. These findings reinforce anaemia as a priority for urgent interventions in pregnancy through pathways independent of lead exposure. However, occupational lead surveillance and control remain important policy considerations.

## Introduction

Anaemia in pregnancy is a major global public health concern, affecting approximately 37% of pregnant women worldwide and substantially increasing the risk of adverse maternal and neonatal outcomes, including preterm delivery, low birth weight, and maternal mortality [1, 2]. Anaemia is disproportionately prevalent in low- and middle-income countries, particularly in sub-Saharan Africa, where complex interactions among nutritional deficiencies, infectious disease, and environmental exposures compound its burden. In a recent systematic review, the prevalence of anaemia among pregnant women in Sub-Saharan Africa was 35.6%, ranging from 7.9 to 76% [3]. In Ghana, data from the 2022 Ghana Demographic and Health Survey revealed that 51.4% of pregnant women were anaemic [4], a prevalence that constitutes a severe public health threat. There exists considerable intra-country variation with anaemia risk being higher among women in the northern part of Ghana compared to those in the south [4]. These geographic disparities reflect differences in socioeconomic status, agricultural practices, access to healthcare, and differential exposure to infectious diseases and environmental toxicants, all of which interact to shape anaemia risk in the Ghanaian context.

Lead (Pb) is a naturally occurring, persistent heavy metal with no known physiological role in the human body. It is classified by the International Agency for Research on Cancer (IARC) as a Group 2A probable human carcinogen [5], and is recognised by major public health authorities as one of the most significant environmental toxicants affecting human health globally [6, 7]. Lead exposure occurs via multiple routes: inhalation of contaminated dust and fumes, ingestion of contaminated food, water, and soil, and, in some contexts, through skin contact. Historically, leaded petrol was the dominant source of environmental lead exposure; however, since the phase-out of leaded gasoline in most sub-Saharan African countries in the early 2000s [8], exposure continues through a diverse array of pathways including informal lead-acid battery recycling, lead-based paint, artisanal and small-scale gold mining (ASGM) activities, leaded cosmetics, piped water, traditional medicines, and the ingestion of clay or soil, a practice known as geophagia that is prevalent among pregnant women in many West African communities [9, 10]. In the specific context of Ghana, ASGM represents a major and growing source of heavy metal contamination of soils, water bodies, and food crops [11, 12]. Lead and other heavy metals, including arsenic and cadmium have been detected at concentrations exceeding guideline values in water samples from ASGM-impacted areas across Ghana, with lead hazard quotients above acceptable threshold of 1.0 documented among mining community residents [13, 14].

The biological plausibility linking lead exposure to anaemia in pregnancy is well established. Over 90% of the lead absorbed by humans is stored in bones and tissues and gets redistributed into blood circulation during pregnancy, lactation and ageing [15–17]. Lead inhibits two critical enzymes in the haem biosynthesis pathway, delta-aminolevulinic acid dehydratase (ALAD) and ferrochelatase, thereby impairing haemoglobin synthesis, reducing red blood cell survival, and ultimately lowering circulating haemoglobin concentrations [18, 19]. Lead also competes with iron for shared transport proteins, including divalent metal transporter-1 (DMT-1) [20], such that iron-deficient individuals absorb lead more readily, creating a synergistic cycle in which pre-existing iron deficiency accelerates lead accumulation during pregnancy. Lead also increases erythrocyte fragility by directly acting on red cell membranes and their enzymes, shortening erythrocyte lifespan and inducing haemolytic anaemia [15, 21]. The combined consequence of impaired haemoglobin synthesis, suppressed red blood cell production, and accelerated erythrocyte destruction produces a characteristic microcytic, hypochromic haemolytic anaemia that is clinically indistinguishable from iron deficiency anaemia without appropriate biochemical investigations. These mechanistic pathways suggest that pregnant women with anaemia or iron deficiency may be particularly vulnerable to the haematological effects of lead, and conversely, that lead exposure may impede the response to standard anaemia treatment [22]. Though some studies have demonstrated associations between even low levels of lead exposure and anaemia among pregnant women [19, 23], other studies did not find any associations [24, 25].

Despite this body of evidence, direct investigation of the relationship between BLLs and anaemia specifically among pregnant women in the Kintampo area of Ghana remains limited. Understanding the trimester-specific distribution of BLLs and the prevalence of anaemia across pregnancy, and the nature of their association is essential for informing targeted clinical and public health responses in this setting. This study therefore sought to determine trimester-specific BLLs and examine their association with anaemia among pregnant women attending antenatal care in the Kintampo area of Ghana.

## Materials and Methods

### Study design and recruitment

This prospective cohort study recruited 300 pregnant women in their first trimester from the Pregnancy Risk, Infant Surveillance, and Measurement Alliance (PRISMA) Maternal and Newborn Health Study, carried out at the Kintampo Health Research Centre (KHRC) between January and November 2024. KHRC is one of six study sites that carried out the PRISMA study. The details of the PRISMA study are explained elsewhere [26, 27]. In brief, the study population included women of reproductive age (15 to 49 years) living within the catchment area of KHRC. For inclusion into the PRISMA, the woman must be less than 20 weeks pregnant (by ultrasound) and willing to deliver and stay up to 6 weeks postpartum in the study area.

### Sample size determination

The sample size was determined using the formula described by Fleiss for cohort studies when comparing two proportions [28]:

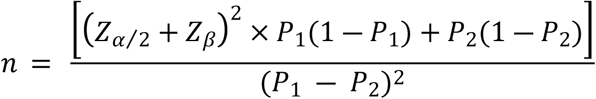

Where:

Zα/2 = 1.96 (95% confidence level)

Zβ = 0.84 (80% power)

P1 = 0.756 (prevalence among exposed group) from a study done in Nigeria [29] since no local prevalence rates were found.

P2 = 0.50 (assumed prevalence among control group)

A total of 104 pregnant women were obtained and this was further increased to 150 to achieve statistical robustness. In order to account for attrition rate of 50%, the sample size was further adjusted to 300 to ensure that a minimum of 150 participants were followed up in the second and third trimesters for analysis.

### Inclusion and exclusion criteria

#### Inclusion criteria

– First trimester pregnant women within the PRISMA cohort as determined by ultrasound (<14 weeks).

#### Exclusion criteria

– Pregnant women on medication for underlying health conditions like high blood pressure, diabetes mellitus, human immunodeficiency virus (HIV) etc., were excluded to prevent the condition or its treatment from influencing the primary outcomes of the study.

### Data collection

#### Sociodemographic and household characteristics

Structured questionnaires were used to collect data on maternal age, parity, marital status, religious affiliation, educational level, employment status, residence type (urban/rural), main household water source, and socioeconomic status measured using wealth quintiles. Parity was categorized as nulliparous, primiparous, and multiparous.

#### Anthropometric measurements

Maternal anthropometric assessments were conducted at enrollment and at all antenatal care (ANC) visits. Height was measured in centimeters using a stadiometer, and weight in kilograms using a calibrated scale. Body mass index (BMI) was computed as weight in kilograms divided by height in meters squared (kg/m²). All measurements were taken using standardized procedures.

### Laboratory measurements

#### Blood sampling

Venous blood was taken from participants in each trimester of pregnancy. Specifically, blood samples were taken at recruitment (<14 weeks) for T1 blood draw, at 20 weeks (±1 week) for T2 and 32 weeks (± 1 week) for T3 blood draw. Participants who missed these scheduled visits had their samples taken whenever they made the visit within the expected window and matched to the specific gestational age. Blood samples were collected into trace element-free ethylenediaminetetraacetic acid (EDTA) tubes for lead estimation and regular EDTA tubes for Full blood count (FBC) for anaemia determination.

#### Haemoglobin measurement to determine Anaemia

Haemoglobin measurement was performed to determine anaemia using a five-part differential haematology analyzer, Sysmex XN 330 (Sysmex Corporation, Japan) at the Seth Owusu Agyei Medical Laboratory of KHRC. Three levels of commercially obtained controls (low, normal and high) were analysed before participant sample analysis. The laboratory participated in external quality assessment provided by the United Kingdom National External Quality Assessment Scheme (UK NEQAS) and the College of American Pathologists (CAP) to monitor the quality of the results generated. Women with anaemia were referred through the routine antenatal care system for further evaluation and management.

#### Blood lead analysis

Blood lead analysis was performed using ICP-MS (NexION 2000, PerkinElmer, USA) at the Chemistry Laboratory of the Ghana Standard Authority (GSA). The GSA is the national statutory body responsible for metrology, standards, testing, inspection and certification. ICP-MS is a highly sensitive analytical method for trace-element quantification, in which the sample is introduced into an argon plasma that atomises and ionises the analyte. The ions generated are then separated on the basis of their mass-to-charge ratio and detected by the mass spectrometer.

An acid diluent was prepared using a 1-L volumetric flask: 7.14 mL of 70% nitric acid, 500 µL of Triton X-100 and 20 mL of methanol were added to approximately 200 mL of distilled water, and the mixture was subsequently brought to a final volume of 1 L with additional distilled water. This yielded a diluent with final concentrations of approximately 0.5% nitric acid, 0.05% Triton X-100 and 2% methanol.

Before analysis, whole-blood specimens were mixed thoroughly to achieve homogeneity. A 0.5-mL aliquot of each blood sample was then transferred into a 25-mL volumetric flask and brought to volume with the acid diluent described above, giving an overall dilution factor of 50-fold. The diluted specimens were analysed on the ICP-MS instrument, and the resulting lead concentrations were corrected for the dilution factor to be reported in µg/dL of whole blood. Calibration and quality-control measures were carried out throughout to maintain analytical accuracy and precision. Given that the study population was unexposed, particular attention was paid to preventing contamination at every stage – specimen handling, reagent preparation and instrumental analysis. Blank samples were run alongside the specimens in each analytical batch to track background lead levels, and rigorous cleaning and rinsing protocols were followed to minimise carry-over contamination between successive samples.

### Statistical analysis

Data were analyzed using STATA 17 software. Continuous variables were summarized using means and standard deviations or geometric means with 95% confidence intervals where distributions were skewed (blood lead levels). Categorical variables were summarized using frequencies and percentages. Anaemia was defined as haemoglobin < 11.0, 10.5 and 11.0g/dL for T1, T2 and T3, respectively [1], while elevated BLL was ≥3.5µg/dL [7]. Differences in blood lead levels across trimesters were assessed using geometric mean ratios (GMRs) with 95% confidence intervals, with T1 as the reference category. Logistic regression models were used to estimate odds ratios (ORs) and 95% confidence intervals for anaemia across trimesters adjusting for covariates and interaction effects. A population-averaged GEE model was used to assess the association between elevated BLL and anaemia across trimesters, adjusting for sociodemographic characteristics.

A two-sided p-value of <0.05 was used as the threshold for statistical significance.

### Ethical considerations

Ethical approval for the conduct of the study was obtained from the Institutional Ethics Committee of the Kintampo Health Research Centre (KHRCIEC/2023-34) and also from the Kwame Nkrumah University of Science and Technology (KNUST) Committee for Human Research Publications and Ethics (CHRPE/AP/1044/23). All participants provided written informed consent prior to recruitment into the study.

## Results

### Participant Characteristics

A total of 300 pregnant women were enrolled during their first trimester (mean gestational age 10.6±2.0 weeks, range 4.9-13.9 weeks) and followed prospectively throughout pregnancy. The cohort retention was 80.3% (241/300) at T2 and 58.3% (175/300) atT3. The primary reason for loss to follow-up was relocation outside the study area mainly during farming and harvesting seasons (Figure 1).

**Fig 1:**
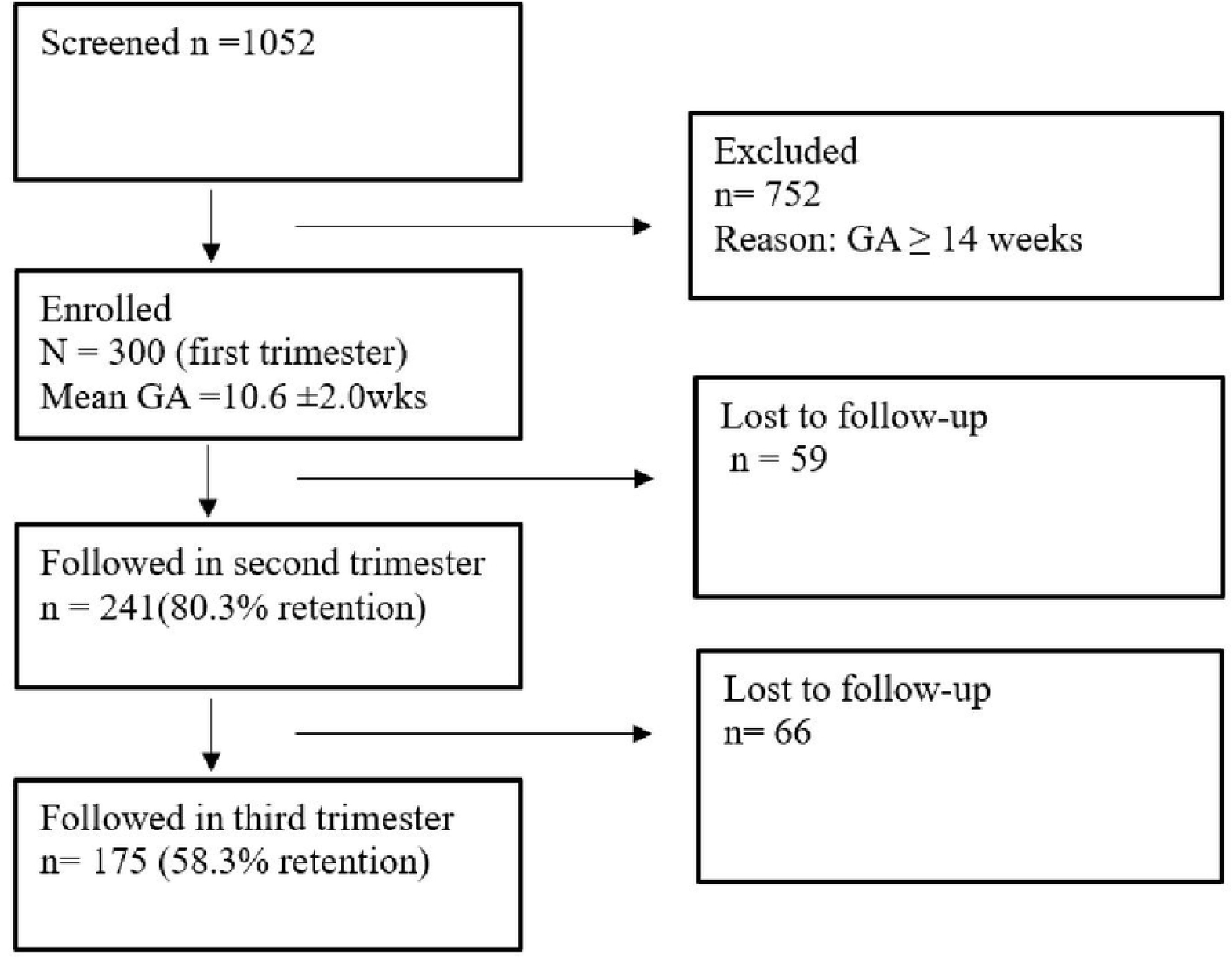
Study flow diagram (GA=gestational age, retention is calculated relative to n=300 at baseline)

The mean age of participants was 27.7±5.8 years (range 18-44years). The majority (81.0%) of participants were between 20-34 years, were nulliparous (82.0%), married (68.0%), Christian (62.0%), and resided in rural areas (70.0%). Over half (57%) had received formal education, with the dominant occupation being farming (41%) or were unemployed (30%). Socioeconomic distribution showed clustering toward lower wealth quintiles, with 55% classified as poor. Mean anthropometric measurements at enrolment were: height 159.3±6.7cm, and weight 59.3±12.4kg (Table 1). Mean gestational age at enrollment was 10.6±2.0 (range 4.9-13.9weeks).

**Table 1:** Demographic characteristics of study participants at recruitment (N=300)

| Characteristic | Enrolment at Gestation < 14 weeks |  |
| --- | --- | --- |
|  | Number (N) | Proportion (%) |
| **Age (yrs)** |  |  |
| < 20 | 17 | 5.7 |
| 20 – 34 | 243 | 81.0 |
| 35 – 49 | 40 | 13.3 |
| **Parity** |  |  |
| Nulliparity | 246 | 82.0 |
| Primiparity | 3 | 1.0 |
| Multiparity | 51 | 17.0 |
| **Marital Status** |  |  |
| Married | 205 | 68.0 |
| Not married | 95 | 32.0 |
| **Religion** |  |  |
| Christian | 186 | 62.0 |
| Muslim | 108 | 36.0 |
| Traditional | 5 | 1.7 |
| Others | 1 | 0.3 |
| **Education** |  |  |
| Formal education | 170 | 56.7 |
| No formal education | 130 | 43.3 |
| **Occupation** |  |  |
| Government worker | 4 | 1.3 |
| Farming | 123 | 41.0 |
| Self-Employed | 41 | 13.7 |
| Artisans | 43 | 14.3 |
| Unemployed | 89 | 29.7 |
| **Residence** |  |  |
| Urban | 91 | 30.3 |
| Rural | 209 | 69.7 |
| **Socioeconomic status** |  |  |
| Poor | 165 | 55.0 |
| Moderate | 104 | 34.7 |
| Rich | 31 | 10.3 |
| **Water Source** |  |  |
| Improved source | 173 | 57.7 |
| Wells | 94 | 31.3 |
| Unsafe source | 33 | 11.0 |
| **Malaria Parasitaemia** |  |  |
| Slide Positive | 10 | 3.3% |
| Slide Negative | 290 | 96.7% |
| **Baseline Anthropometry** |  |  |
| Height (cm) | 159.3 ± 6.7 (range 109.0 – 177.0) |  |
| Weight (kg) | 59.3 ± 12.4 (range 39.0 – 157.0) |  |
| Body Mass Index (BMI) | 23.0 ± 3.8 (range 16.7 – 44.4) |  |

### Blood Lead Levels

Blood lead levels increased progressively across trimesters. The geometric mean BLL was 1.0 µg/dL (95% CI: 0.90-1.12) in T1, rising to 1.3 µg/dL (95% CI: 1.15-1.42) in T2 and 1.5 µg/dL (95% CI: 1.28-1.75) in T3. Compared to T1, the geometric mean ratio (GMR) was 1.3 (95% CI: 1.08-1.49, *p*=0.003) for T2 and 1.5 (95% CI: 1.25-1.76, *p*<0.001) for T3, indicating significant increases with increasing trimesters (Table 2).

**Table 2:**
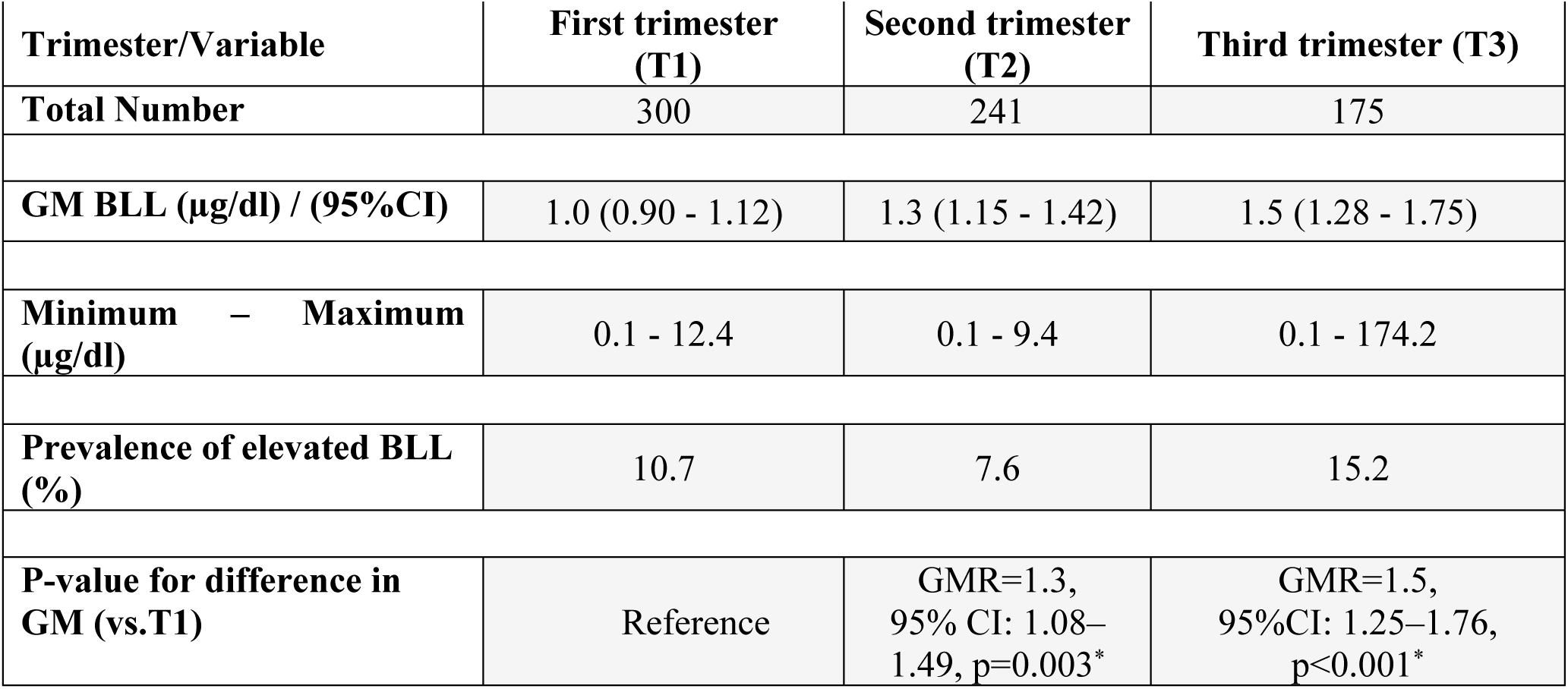
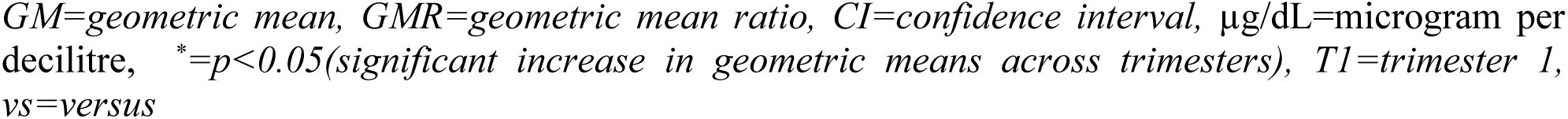
Blood Lead Levels in Pregnant Women by Trimester.

| Trimester/Variable | First trimester (T1) | Second trimester (T2) | Third trimester (T3) |
| --- | --- | --- | --- |
| **Total Number** | 300 | 241 | 175 |
| **GM BLL (µg/dl) / (95%CI)** | 1.0 (0.90 - 1.12) | 1.3 (1.15 - 1.42) | 1.5 (1.28 - 1.75) |
| **Minimum – Maximum (µg/dl)** | 0.1 - 12.4 | 0.1 - 9.4 | 0.1 - 174.2 |
| **Prevalence of elevated BLL (%)** | 10.7 | 7.6 | 15.2 |
| **P-value for difference in GM (vs.T1)** | Reference | GMR=1.3, 95% CI: 1.08–1.49, $p=0.003^*$ | GMR=1.5, 95%CI: 1.25–1.76, $p<0.001^*$ |

*GM=geometric mean, GMR=geometric mean ratio, CI=confidence interval, µg/dL=microgram per* *decilitre, \*= $p<0.05$ (significant increase in geometric means across trimesters), T1=trimester 1,* *vs=versus*

The prevalence of elevated BLL (≥3.5 µg/dL) was 10.7% in T1, 7.6% in T2, and 15.2% in T3. While the overall range of BLL was 0.1-12.4 µg/dL in T1, 0.1-9.4 µg/dL in T2, and T3 showed substantial variability, with one extreme outlier at 174.2 µg/dL (range 0.1-174.2 µg/dL).

### Anaemia Prevalence in Pregnancy

Mean haemoglobin concentration declined progressively throughout pregnancy, from 11.2±1.1 g/dL in T1 to 10.3±1.0 g/dL in T2 and 10.2±1.0 g/dL in T3. Similarly, anaemia prevalence (Hb <11 g/dL in T1 and T3 and <10.5g/dL in T2) increased substantially from 34.7% in T1 to 59.3% in T2 and 76.0% in T3 (Table 3).

**Table 3:** Haemoglobin levels and prevalence of anaemia among pregnant women.

| Trimester/Variable | T1 | T2 | T3 |
| --- | --- | --- | --- |
| Total Number | 300 | 241 | 175 |
| Mean Hb (g/dL) (SD) | 11.2 ± 1.1 | 10.3±1.0 | 10.2±1.0 |
| Minimum – Maximum (g/dL) | 6.7 – 15.7 | 6.9 – 13.8 | 7.1 – 13.6 |
| Prevalence of Anaemia (%) | 34.7 | 59.3 | 76.0 |
| Odds of Anaemia (vs. T1) | Reference | (OR = 2.78, 95% CI: 2.06–3.74, $p<0.001$) | (OR = 6.38, 95% CI: 4.41–9.24, $p<0.001$) |

T1=first trimester, T2=second trimester, T3=third trimester, Hb=haemoglobin, g/dL=grams per decilitre, SD=standard deviation, OR=odds ratio, CI=confidence interval, vs=versus, $p<0.05$ =significant

Compared to T1, the odds of anaemia were significantly higher in both T2 (OR=2.78, 95% CI: 2.06-3.74, *p*<0.001) and T3 (OR=6.38, 95% CI: 4.41-9.24, *p*<0.001), indicating a strong temporal pattern of increasing anaemia development across pregnancy (Table 3).

### Association between Elevated BLL and Anaemia in Pregnancy

Cross-tabulations of elevated BLL (≥3.5 µg/dL) against anaemia across all three trimesters are presented in Table 4. Across all time points, anaemia was more prevalent among women with elevated BLL than those with normal BLL, although none of the associations reached statistical significance.

**Table 4:** Cross-tabulations of Elevated Blood Lead Levels and Anaemia by Trimester.

| BLL Group | No Anaemia<br>n (%) | Anaemia<br>n (%) | Total<br>(N) | Odds Ratio<br>(95% CI) | P-value |
| --- | --- | --- | --- | --- | --- |
| T1(n=300): Anaemia threshold: Hb < 11.0 g/dL |  |  |  |  |  |
| Elevated BLL (≥3.5 μg/dL) | 18 (56.3%) | 14 (43.8%) | 32 | 1.54 (0.73–3.23) | 0.25 |
| Normal BLL | 178 (66.4%) | 90 (33.6%) | 268 | 1.00 (Ref) |  |
| Total | 196 (65.3%) | 104 (34.7%) | 300 | NA |  |
| T2 (n = 241): Anaemia threshold: Hb < 10.5 g/dL |  |  |  |  |  |
| Elevated BLL ($\geq 3.5$ $\mu\text{g/dL}$) | 7 (36.8%) | 12 (63.2%) | 19 | 1.19 (0.45–3.14) | 0.81 |
| Normal BLL ($< 3.5$ $\mu\text{g/dL}$) | 91 (41.0%) | 131 (59.0%) | 222 | 1.00 (Ref) | |
| **Total** | 98 (40.7%) | 143 (59.3%) | 241 | NA |  |
| **T3 (n = 175): Anaemia threshold: Hb < 11.0 g/dL** |  |  |  |  |  |
| Elevated BLL ($\geq 3.5$ $\mu\text{g/dL}$) | 6 (23.1%) | 20 (76.9%) | 26 | 1.06 (0.40–2.85) | 0.91 |
| Normal BLL ($< 3.5$ $\mu\text{g/dL}$) | 36 (24.2%) | 113 (75.8%) | 149 | 1.00 (Ref) | |
| **Total** | 42 (24.0%) | 133 (76.0%) | 175 | NA |  |

*T1=first trimester, T2=second trimester, T3=third trimester, n=number per category, N=total number per category, Hb=haemoglobin, g/dL=grams per decilitre, OR=odds ratio, CI=confidence interval, NA=not applicable, $p<0.05$ =significant, $\mu\text{g/dL}$ =microgram per decilitre.*

Trimester-specific multivariable logistic regression models were employed to identify independent predictors of maternal anaemia across T1, T2, and T3 of pregnancy. Across all trimesters, elevated BLLs were not independently associated with maternal anaemia in either the crude or adjusted analyses [adjusted for maternal age, parity, education, malaria parasitaemia, body-mass index (BMI), gestational age, occupation], suggesting that lead exposure, at the levels observed in this cohort, may not be a primary determinant of anaemia risk during pregnancy (Table 5).

**Table 5:**
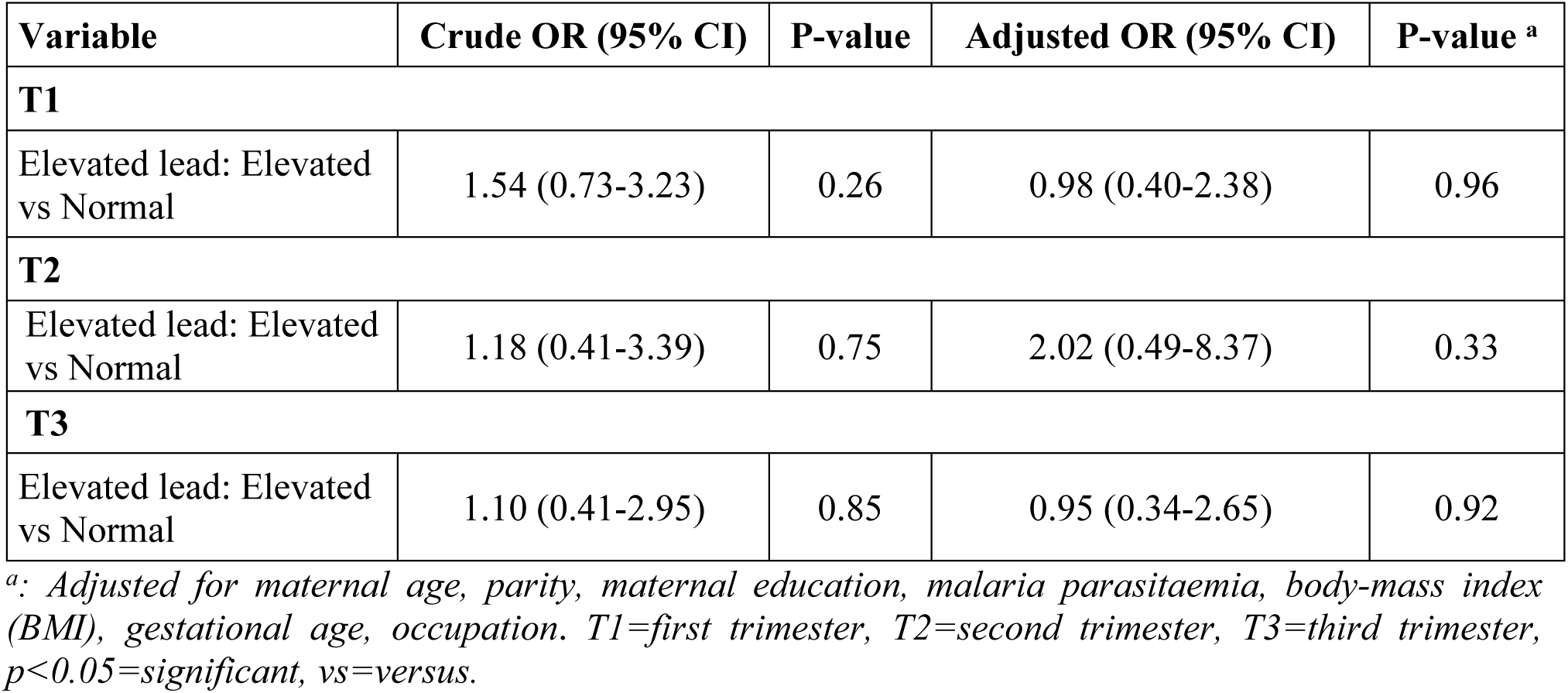
Logistic regression to determine association between elevated BLL and Anaemia.

A population-averaged GEE model was used to examine the association between elevated BLL and anaemia across trimesters. Elevated BLL was associated with higher odds of anaemia (OR = 1.29, 95% CI: 0.77–2.17, p = 0.327), though this association was not statistically significant, and the wide confidence interval indicates considerable uncertainty around the estimate.

## Discussion

In this prospective, trimester-specific cohort of pregnant women in the Kintampo area of Ghana, BLLs increased significantly across gestation, from a geometric mean of 1.0 µg/dL in the T1 to 1.5 µg/dL in T3, while anaemia prevalence more than doubled, from 34.7% to 76.0% respectively. Additionally, elevated BLL suggested a trend toward increased odds of anaemia across trimesters (OR = 1.29, 95% CI: 0.77–2.17).

Despite this increase and the parallel escalation of BLL across trimesters, and despite a well-established biological rationale for a causal relationship between the two, elevated BLL (≥3.5 µg/dL) was not independently associated with anaemia at any trimester after adjustment for maternal age, parity, education, malaria parasitaemia, body mass index, gestational age, and occupation. These findings are consistent with other studies that found no significant associations in lead levels and anaemia or haemoglobin among pregnant women [24, 25].

The progressive rise in BLL across pregnancy supports existing knowledge that skeletal calcium turnover mobilizes stored lead in late pregnancy [16, 17, 30]. This is consistent with isotope-based evidence that the maternal skeleton becomes an increasingly important source of circulating lead as fetal calcium demand rises: the cumulative increase in BLL relative to the T1 was larger by T3 (GMR 1.49, 95% CI 1.25-1.76) than by T2 (GMR 1.27, 95% CI 1.08-1.49), consistent with accelerating mobilisation as pregnancy advances[15–17].

Despite this upward trajectory, absolute BLLs were low overall by international standards. Geometric means of 1.0-1.5 µg/dL fall below the CDC’s level of concern (≥3.5 µg/dL) [7]. Lead levels recorded in this study were still lower than national pooled means reported for children in a systematic review spanning 34 low-and middle-income countries, where country-level averages ranged from 1.7 µg/dL in Ethiopia to 9.3 µg/dL in Palestine, though adult values ranged from 0.4 in Sudan to 11.4 in Pakistan [8]. Similarly, mean blood lead levels in pregnant women were 2.8 in Mexico [31], 8.1 in India [19], 12.1 in Jordan [25], 36.4 in Nigeria [32], much higher than observed in this cohort. This is reassuring at the population level, since very elevated BLLs are associated with adverse pregnancy outcomes, though still cautious with the knowledge that there are no known safe levels of blood lead [6, 7]. Documented sources of lead exposures in Ghana include informal artisanal and small-scale gold mining [12, 14] and geophagia, a culturally embedded practice among pregnant Ghanaian women shown to carry a lead-related target hazard quotient above 1.0 in clay sampled in the Volta Region [10]. The picture may therefore be different from elsewhere in Ghana where there is mining and the potential for lead to be released from artisanal mining activities.

The high burden of anaemia observed in this study (34.7% in T1, 59.3% in T2 and 76.0% in T3) is consistent with the 2022 Ghana Demographic and Health Survey estimates of 51.4% anaemia prevalence among pregnant women nationally, a figure sitting between our T1 and T2 estimates [4]. A T3 prevalence of 76.0% is nevertheless high even by sub-Saharan African standards and confirms that anaemia, irrespective of any association with lead, remains the dominant and most actionable maternal morbidity in this population, with well-documented downstream adverse consequences such as preterm birth, low birth weight, and maternal mortality [2].

The absence of an independent BLL association with anaemia in pregnancy seems to contradict existing evidence that lead suppresses erythropoietin production and competes with iron for shared intestinal transporters such as DMT-1, compounding the haem-synthesis inhibition (via ALAD and ferrochelatase) [15, 20]. Several features of our study help explain rather than contradict this literature. Most women in our study had BLLs well below the range in which haematological effects have been reported [33]. This may explain why no significant association was found between BLLs and anaemia in this cohort.

### Strengths and limitations of this study

This study’s principal strength is its prospective, trimester-specific design, which is uncommon in the sub-Saharan African literature on gestational lead exposure, and therefore allows for characterisation of trimester-specific estimates rather than a single cross-sectional snapshot. To the best of our knowledge, this is the first study that has measured blood lead levels in pregnant women in Ghana and to further determine associations with anaemia. Additionally, BLL was quantified by ICP-MS, the gold standard method, with the analysis performed at the reputable ISO certified Ghana Standard Authority Laboratory. Recruitment through the multi-country PRISMA platform means data collection followed a harmonised protocol amenable to future pooled analysis alongside the study’s other sites in India, Pakistan, Kenya, and Zambia [26].

The limitation of this study was its inability to measure bone lead stores which estimate total body burden of lead.

## Conclusion

Pregnant women attending antenatal care in the Kintampo area experience a rising and substantial burden of anaemia across gestations, but this study did not find evidence that BLL was independently linked to anaemia in pregnancy. Anaemia management therefore remains an urgent, largely lead-independent clinical priority in this setting, while continued surveillance and source control of lead exposure, particularly around artisanal mining and geophagia remain necessary. Larger, adequately powered studies combining trimester-specific biomonitoring with individual-level attribution of environmental exposure sources are needed to determine whether associations observed in more heavily exposed settings are also present in the Ghanaian population.

## Author contributions

**Conceptualization:** Veronica Agyemang, Otchere Addai Mensah, Kwaku Poku Asante **Data curation**: Veronica Agyemang, Dennis Gyasi Konadu, Eliezar Ofori-Lartey, Stephaney Gyaase **Formal analysis**: Veronica Agyemang, Joana Edem Koto, Stephaney Gyaase **Funding acquisition**: Kwaku Poku Asante, Sam Kofi Newton, Charlotte Tawiah Agyemang, Emily R. Smith, Veronica Agyemang **Methodology**: Veronica Agyemang, Dennis Gyasi Konadu, David Dosoo, Dennis Adu-Gyasi **Project administration**: Charlotte Tawiah Agyemang, Veronica Agyemang, Irene Apewe Adei, Lawrence Febir **supervision**: Otchere Addai Mensah, Lilian Antwi Boateng, Benedict Sackey Kwaku Poku Asante **Writing original draft:** Veronica Agyemang **Writing** – review and editing: All authors except Charlotte Tawiah Agyemang (Blessed memory).

## Data Availability

The analytical datasets generated and analysed during this study are not publicly available but may be made available upon reasonable request, subject to approval by the principal investigators and relevant ethical committees.

## Acknowledgement

The authors are grateful to the Director, Management and Staff of the Kintampo Health Research Centre, especially the Laboratory, Field and Data teams for the immense support received during the conduct of this study. Also grateful to Dr. Samuel Ekow Bernard Harrison for his valuable support during data analysis and to Mr. Prosper Mwintuuro, Mr. Love Ankra, Ms. Mavis Ntibea, Ms. Mary Abapa, Ms. Leticia Fosuah, for their support in laboratory procedures. We also thank the team at the Ghana Standards Authority led by Dr. Paul Fosu for their support with blood lead analysis. Our appreciation also goes to all the pregnant women who gave their consent to be part of this study. This study also received support from the Gates Foundation through grants received by the Kintampo Health Research Centre [grant number: INV-002220 and INV-037626 to KPA, CTA, and SN.

